# Disengagement from care and disease severity among people self-testing positive for hepatitis C in Nigeria, Cameroon, and South Africa: a multi-country cohort analysis of implementation studies

**DOI:** 10.64898/2026.02.20.26346699

**Authors:** Yasmin Dunkley, Bernhard Kerschberger, Victor Adepoju, Hedgar Plessy Mboussam, Vanessa Msolomba, Mohammed Majam, Annie Michele Mabally, Adetiloye Oniyire, Augustin Choko, Pitchaya Indravudh, Nicola Desmond, Peter MacPherson, Elizabeth Lucy Corbett, Karin Hatzold

## Abstract

**Introduction:** Access to Hepatitis C virus (HCV) testing and treatment remains low globally. HCV self-testing (HCVST) may facilitate diagnosis and cure. We analysed treatment uptake and outcomes following a positive HCVST result in three distinct African epidemic contexts.

**Methods:** A multi-country cohort study nested within HCVST implementation programmes in Cameroon, Nigeria, and South Africa (May 2023–May 2024). Adults (≥18 years) with positive HCVST results were followed through confirmatory testing and care outcomes until last event.

Co-primary outcomes were: (i) cascade progression, (treatment initiation and sustained virological response [SVR]); estimated using country-cascades; and (ii) cumulative incidence of disengagement from care, estimated using Bayesian competing-risks survival models. Analyses were conducted separately for South Africa and jointly for Cameroon–Nigeria due to structural differences in service organisation. Covariate associations were estimated as hazard ratios. Disease severity was assessed through fibrosis staging derived from AST-to-platelet ratio index (APRI).

**Results:** 1,341 participants had positive HCVST results (117 in Cameroon, 226 in Nigeria, 998 in South Africa). Among laboratory confirmed HCV cases, treatment initiation and SVR were highest in Cameroon (Tx 98.6%, 71/72; SVR 96.4%, 53/55), followed by Nigeria (Tx 90.8%, 168/185; SVR 91.8%, 56/62), and low in South Africa (Tx 4.3%, 37/854; SVR 60.6%, 3/5).

Crude disengagement was lowest in Nigeria (24.4%; 95% CrI 19.1%-30.3%), followed by Cameroon (52.4%; 95% CrI 44.4%-61.2%), and South Africa (77.9%; 95% CrI 76.2%-79.8%). By 24-weeks, disengagement was lower in specialist hospitals than community sites in Cameroon and Nigeria. In South Africa, the greatest predictor of disengagement was HIV positive status (HR 1.96; 95% CrI 1.71 to 2.23).

Viraemia exceeded regional estimates (82.2%, 1102/1341), with liver scarring highest in Cameroon (fibrosis: 8.3%, cirrhosis: 6.9%) and lowest in South Africa (2.9% and 1.6%, respectively).

**Conclusion:** HCV self-testing enabled detection of HCV cases, including severe disease, but poorer progression in community settings suggests decentralised treatment pathways require strengthening to realise cure.

## Introduction

An estimated 50 million people are living with hepatitis C virus (HCV) [1]. HCV disease burden is particularly pronounced across Africa [2, 3], with recent general population seroprevalence estimates at 2.3% [3]. The region has approximately 8 million people living with HCV of which only 13% have been diagnosed, and 3% initiated treatment [1]. This is despite the release in 2013 of second generation Direct Acting Antivirals (DAAs) [4], which can cure more than 95% of people living with HCV after a 12-week course [5]. Despite some countries, such as Egypt, having made remarkable progress towards HCV elimination [6], in much of the rest of the continent, HCV care and prevention programs may have supportive national policy but limited implementation [7]. For the African continent, scaling up access to viral hepatitis testing is considered the crucial entry point to care. Yet, limited decentralization of healthcare services generally, as well as traditional models of diagnostic testing characterized by centralized laboratory systems have hindered testing and treatment scale-up [8]. This challenge is exacerbated by overburdened health systems [1, 7].

HCV self-testing (HCVST) is a WHO-endorsed strategy to expand access to HCV testing as a complement to facility-based testing [9], with impact contingent on effective linkage and retention across the diagnostic and treatment cascade. This builds on the highly successful experience of HIV self-testing [1, 10]. However, the implementation of HIV self-testing adopted in 2016 was against a backdrop of comparatively well-established prevention and treatment programmes – as a “last mile” solution to epidemic end [11]. HCV self-testing is promoted against a backdrop of limited existing HCV care and prevention programming. In recognition of this, WHO stresses the importance of integrating HCV services with other services, such as within harm-reduction and HIV services as part of simplified service delivery models [10, 12]. There is also a clear resource rationale for integration when considering the client populations reached within harm reduction and HIV programmes are likely those most at risk of HCV [10]. Yet, even with the example of HIV self-testing, linkage to confirmatory testing and ART initiation has ranged from 23–68% [13]. Established barriers to HIV treatment include that optimal systems for linking clients to HIV care are not well established [13]. The introduction of HCV self-testing in emerging care and prevention programs in resource-limited settings raises questions about its effectiveness and sustainability in identifying and retaining patients.

To answer these questions, we investigated disengagement from care across cascades and disease severity outcomes among people testing positive with HCVST in Nigeria, Cameroon, and South Africa; African countries conducting parallel HCVST implementation studies under the multi-country STAR initiative [14–16]. Studies were implemented independently but used comparable self-testing products and cascade definitions. These three different African countries represent distinct epidemiological contexts, populations at risk and delivery models. These differences enable a natural comparison to investigate linkage across different implementation contexts in overburdened health systems. This study aimed to: (i) assess disengagement from care across cascades in three contrasting African epidemic contexts; and (ii) describe the disease severity profile of those reached through HCVST.

## Methods

### Ethics statement

Individual studies and cross-country analyses were approved at three-levels nationally, through parent research agencies and through the Ethics Research Committee of the World Health Organization (WHO ERC). In Nigeria, an exemption for state-level ethics review was also obtained. Details of all approvals can be found in (**S1)**.

Written informed consent was obtained from all participants.

### Study design

We conducted a multi-country cohort study nested within HCV self-testing implementation studies in Nigeria, Cameroon, and South Africa [17–19]. We followed participants with positive HCVST results from first positive result to downstream HCV care outcomes. Reporting follows the STROBE guidelines for cohort studies [20].

### Implementation overview

Between 30^th^ May 2023 and 28^th^ May 2024, decentralised HCV self-testing (HCVST) interventions were implemented in Nigeria, Cameroon, and South Africa. Across all three countries, WHO-pre-qualified HCVST kits were used (OraQuick oral-fluid, OraSure Technologies Inc., USA; and First Response blood-based HCV Card Test, Premier Medical Corps Ltd., India). Assisted or unassisted testing was provided at study sites. Sites in each country were purposively selected in collaboration with implementing partners and national programmes as part of the ongoing HCVST implementation studies. Sites were chosen based on their ability to reach populations at elevated risk of HCV through existing service platforms (e.g., harm reduction, key population services, outpatient facilities) and their operational capacity to deliver confirmatory testing and treatment. Positive HCVST results were linked to confirmatory HCV RNA PCR testing either onsite or near-site, and participants with viraemia were assessed for treatment eligibility and supported to initiate free direct-acting antivirals (DAAs, sofosbuvir/velpatasvir), through national programmes or study-linked provision. Cascade outcomes - self-test positivity, confirmatory testing, treatment initiation, treatment completion and sustained virological response - were defined in a harmonised manner across protocols, allowing cross-country analysis despite contextual variation in models of care.

### Models of care

Although interventions shared common products and cascade definitions, each country implemented different models of care reflecting different epidemic burden, risk groups, and health systems (**Table 1**):

**Table 1.**
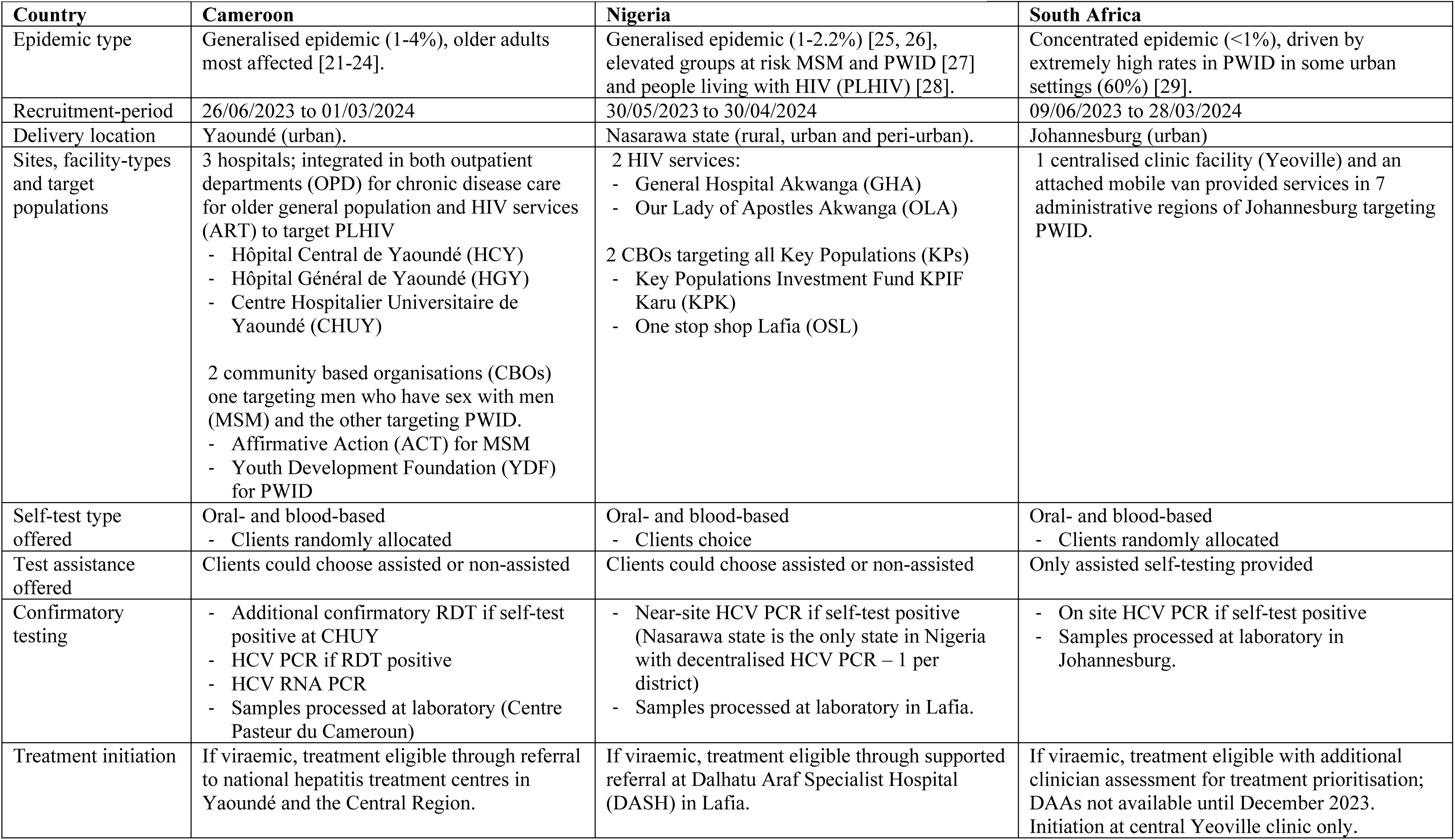
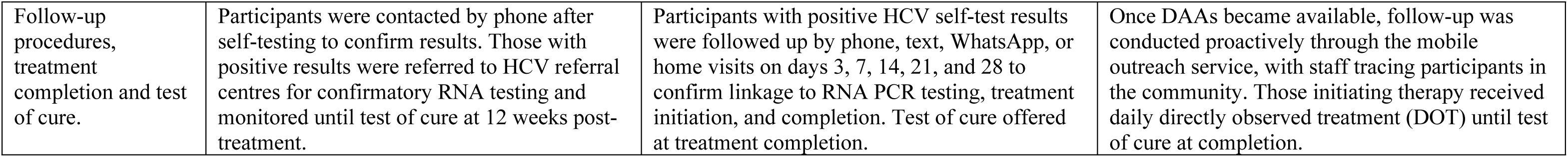
Summary of key characteristics of HCV-diagnostic models of care in each country.

| Country | Cameroon | Nigeria | South Africa |
| --- | --- | --- | --- |
| Epidemic type | Generalised epidemic (1-4%), older adults most affected [21-24]. | Generalised epidemic (1-2.2%) [25, 26], elevated groups at risk MSM and PWID [27] and people living with HIV (PLHIV) [28]. | Concentrated epidemic (<1%), driven by extremely high rates in PWID in some urban settings (60%) [29]. |
| Recruitment-period | 26/06/2023 to 01/03/2024 | 30/05/2023 to 30/04/2024 | 09/06/2023 to 28/03/2024 |
| Delivery location | Yaoundé (urban). | Nasarawa state (rural, urban and peri-urban). | Johannesburg (urban) |
| Sites, facility-types and target populations | 3 hospitals; integrated in both outpatient departments (OPD) for chronic disease care for older general population and HIV services (ART) to target PLHIV<br>- Hôpital Central de Yaoundé (HCY)<br>- Hôpital Général de Yaoundé (HGY)<br>- Centre Hospitalier Universitaire de Yaoundé (CHUY)<br>2 community based organisations (CBOs) one targeting men who have sex with men (MSM) and the other targeting PWID.<br>- Affirmative Action (ACT) for MSM<br>- Youth Development Foundation (YDF) for PWID | 2 HIV services:<br>- General Hospital Akwanga (GHA)<br>- Our Lady of Apostles Akwanga (OLA)<br>2 CBOs targeting all Key Populations (KPs)<br>- Key Populations Investment Fund KPIF Karu (KPK)<br>- One stop shop Lafia (OSL) | 1 centralised clinic facility (Yeoville) and an attached mobile van provided services in 7 administrative regions of Johannesburg targeting PWID. |
| Self-test type offered | Oral- and blood-based<br>- Clients randomly allocated | Oral- and blood-based<br>- Clients choice | Oral- and blood-based<br>- Clients randomly allocated |
| Test assistance offered | Clients could choose assisted or non-assisted | Clients could choose assisted or non-assisted | Only assisted self-testing provided |
| Confirmatory testing | - Additional confirmatory RDT if self-test positive at CHUY<br>- HCV PCR if RDT positive<br>- HCV RNA PCR<br>- Samples processed at laboratory (Centre Pasteur du Cameroun) | - Near-site HCV PCR if self-test positive (Nasarawa state is the only state in Nigeria with decentralised HCV PCR – 1 per district)<br>- Samples processed at laboratory in Lafia. | - On site HCV PCR if self-test positive<br>- Samples processed at laboratory in Johannesburg. |
| Treatment initiation | If viraemic, treatment eligible through referral to national hepatitis treatment centres in Yaoundé and the Central Region. | If viraemic, treatment eligible through supported referral at Dalhatu Araf Specialist Hospital (DASH) in Lafia. | If viraemic, treatment eligible with additional clinician assessment for treatment prioritisation; DAAs not available until December 2023. Initiation at central Yeoville clinic only. |
| Follow-up procedures, treatment completion and test of cure. | Participants were contacted by phone after self-testing to confirm results. Those with positive results were referred to HCV referral centres for confirmatory RNA testing and monitored until test of cure at 12 weeks post-treatment. | Participants with positive HCV self-test results were followed up by phone, text, WhatsApp, or home visits on days 3, 7, 14, 21, and 28 to confirm linkage to RNA PCR testing, treatment initiation, and completion. Test of cure offered at treatment completion. | Once DAAs became available, follow-up was conducted proactively through the mobile outreach service, with staff tracing participants in the community. Those initiating therapy received daily directly observed treatment (DOT) until test of cure at completion. |

### Participants

In Nigeria and South Africa, any adult aged ≥18 years attending one of the study sites was eligible for HCV self-testing. In Cameroon, any adult aged ≥21 years attending one of the study sites was eligible, except at the OPD chronic disease clinic where recruitment was restricted to adults aged ≥45 years. Individuals with known prior HCV diagnosis or prior completion of HCV treatment were excluded from participation. Recruitment was pragmatic and opportunistic mirroring real-life implementation. For this cross-country analysis, the study population comprised all participants who received a positive HCV self-test result within the study window.

### Outcome variables

The study had two co-primary outcomes. The first was successful cascade progression, defined in line with the WHO 2024 consolidated guidelines on person-centred viral hepatitis strategic information [30]. For each country, successful progression was defined in two ways: (i) treatment initiation among participants with confirmed viraemia; and (ii) sustained virologic response (SVR) among participants with a documented treatment outcome by the end of the observation period. The observation period ended after the max-time in days until the last recorded event outcome per participant; Cameroon (272 days), Nigeria (239 days) and South Africa (329 days).

The second co-primary outcome was the cumulative incidence of disengagement from care estimated by country using a competing-risks framework. Because date information were not consistently available for all cascade steps across countries, time-to-event analyses were anchored to four milestone dates that were reliably recorded: date of (i) HCV self-test, (ii) Confirmatory PCR, (iii) Treatment initiation, and (iv) Treatment completion. Programmatic data did not include explicit dates of disengagement. Disengagement was therefore inferred as non-progression to the next milestone (i.e., missing outcome data) assigned at the last recorded event date for the relevant step. Follow-up ended at the earliest of inferred disengagement or a competing event that precluded further cascade progression (i.e., became ineligible or died). The database was closed at the last recorded event date.

The secondary outcome was disease severity. Virological status was reported as HCV RNA detectable or undetectable aligning with WHO treatment recommendations [31, 32]. The AST-to-platelet ratio index (APRI) was assessed in a subset of participants with viraemia; fibrosis staging was derived from APRI thresholds according to WHO recommendations (APRI >1.5 was considered significant fibrosis, of which APRI >2.0; cirrhosis [31]). Participants with viraemia without staging data were classified as unknown.

All outcome data was derived from study records and/or abstracted from patient’s clinical files.

### Exposure variables

Sociodemographic characteristics collected consistently across all three-sites were age, sex, and education,. Age was grouped into 18–24, 25–34, 35–44, 45–54, and 55+ years reflecting data capture in country. Education was harmonised for cross-country analysis into completion of primary or less, secondary, and tertiary.

Key population identity (KP) was recorded differently by site. To maximise comparability, a binary variable (any KP identity yes/no) was generated. Country-specific definitions differed as follows: in Nigeria, KP included PWID, men who have sex with men (MSM), sex workers (SW), and people who had ever paid for sex; in Cameroon, KP was restricted to PWID and MSM; in South Africa, KP included PWID, MSM and SW, and a new category people who use drugs (PWUD). Only one KP identity was recorded per participant, limiting multi-category analyses. The exception was PWID, which was consistently measured across all three sites using the same question: *“Have you ever injected drugs for non-medical reasons?”* HIV status was inferred by ART site attendance in Cameroon. In Nigeria, HIV status was self-reported and confirmed with clinical records where possible. In South Africa, HIV status was captured directly through programmatic data abstraction, as study participants were also engaged in a harm reduction programme providing routine HIV testing services.

HCV testing-related exposures were site type, mode of assistance, and self-test type. Site type was harmonised as: HIV/ART clinic; outpatient department (OPD); or community-based organisation (CBO). Across all countries, CBO sites were designed to provide care to KPs. Mode of assistance was categorised as assisted versus unassisted HCV self-testing, and test type was classified as oral fluid versus blood-based kit.

### Statistical methods

All analyses were performed in R version 4.5.0 (R Foundation for Statistical Computing, Vienna, Austria [33]) using the using the brms [34], posterior [35], and cmdstanr [36] packages.

Descriptive statistics were used to characterise sociodemographic and testing characteristics by country. Categorical variables were summarised as frequencies and proportions and compared using chi-square tests.

For the first co-primary outcome (successful cascade progression), the two WHO milestone indicators for successful cascade progression were compared against the country-level HCV care cascades. Cascade steps were i) HCV self-test positive (entry point into the cohort) ii) confirmatory HCV RNA PCR conducted, iii) viral load result received, iv) confirmed viraemia, v) treatment initiated, vi) treatment completed, vii) treatment outcome recorded, and viii) sustained virologic response (SVR) achieved. At each cascade-step, participants were categorised as either: engaged, ineligible to progress along the cascade (competing risk) or disengaged (missed/never completed the step despite eligibility). Proportions were reported using restricted denominators i.e., only of those eligible for that step. Cascade drop-offs and 95% confidence intervals were calculated by country.

For the second co-primary outcome (cumulative incidence of disengagement), we modelled accelerated failure time using Bayesian Weibull regression with a competing-risks framework. Analyses were conducted separately for South Africa and for a combined Cameroon–Nigeria dataset because of structural differences in service organisation. Unadjusted models were fitted to obtain country-level cumulative incidence functions for disengagement, summarised from the posterior predictive distribution at fixed timepoints (4-, 12-, 24- weeks, and the end of the country-specific observation period; medians with 95% credible intervals). We used weakly informative priors centred at no effect for regression coefficients and weakly regularising priors for the standard deviations of site-level random effects. Model convergence was assessed by inspecting traceplots, by Gelman-Rubin statistics, and by parameter effective sample sizes.

Hazard ratios (HRs) for disengagement, and their corresponding 95% credible intervals were obtained by exponentiating 4000 posterior samples of the relevant regression coefficients from adjusted models. Hazard ratios were used to explore whether individual demographic or behavioural characteristics were associated with disengagement; site was included as a random intercept in all adjusted models to capture between-site heterogeneity. In the Cameroon–Nigeria model, adjustment covariates were age, sex, education, test type, mode of assistance, key population identity and country. HIV status was not included due to the extent of unknown data in Cameroon. In South Africa, where all participants were recruited through the same harm-reduction service, all identified as key populations, and all received assisted self-testing, covariate adjustment was restricted to age, sex, education, HIV status and test type. Facility type was not included as a predictor because of structural differences in facility-types between countries, and lack of within-country variation in South Africa.

Given high site-heterogeneity, we derived site-specific posterior predictive probabilities of disengagement at day-180 and displayed these in forest plots to investigate residual heterogeneity not explained by individual-level covariates. Case-complete analyses were conducted; there were n=11 missing from the pooled Cameroon-Nigeria model (N=329), and none missing from the South African model (N=998).

The secondary outcome (disease severity) was described using frequencies and proportions by country with chi-square tests.

## Results

### Participant throughput and characteristics

1341 participants tested positive using HCVST kits during the study period of a total pooled participant pool of 6,205 (overall yield; 21.6%, 95% CI 20.6% to 22.7%). Almost three-quarters of the positive participants included in this study came from South Africa (74.4%, 998/1,341), followed by Nigeria (16.9%, 226/1341) and Cameroon (8.7%, 117/1341). This was the inverse of the numbers tested in each country setting (South Africa N=1566, Nigeria, N=1995 and Cameroon, N=2664) reflecting differences in diagnostic yield across settings.

Of the positive participants, there were substantial differences in socio-demographic characteristics between countries (**Table 2**), in particular between age, HIV status and Key Population identity.

**Table 2.** Sociodemographic and testing characteristics of people testing HCV positive using HCVST in Cameroon, Nigeria and South Africa.

| Indicator* | Total<br>N = 1,341<br>(%) | Cameroon<br>n = 117<br>(%) | Nigeria<br>n = 226<br>(%) | RSA<br>n = 998<br>(%) | p-value<sup>†</sup> |
| --- | --- | --- | --- | --- | --- |
| Sociodemographic characteristics |  |  |  |  |  |
| Gender |  |  |  |  |  |
| Female | 279 (20.8%) | 63 (53.8%) | 139 (61.5%) | 77 (7.7%) | <0.001 |
| Male | 1062 (79.2%) | 54 (46.2%) | 87 (38.5%) | 921 (92.3%) |  |
| Age group<sup>1</sup> |  |  |  |  |  |
| 18-24 | 75 (5.6%) | 6 (5.1%) | 7 (3.1%) | 62 (6.2%) | <0.001<sup>1</sup> |
| 25-34 | 745 (55.6%) | 9 (7.7%) | 44 (19.5%) | 692 (69.3%) |  |
| 35-44 | 302 (22.5%) | 6 (5.1%) | 67 (29.6%) | 229 (22.9%) |  |
| 45-54 | 91 (6.8%) | 12 (10.3%) | 65 (28.8%) | 14 (1.4%) |  |
| ≥55 | 117 (8.7%) | 84 (71.8%) | 32 (14.2%) | 1 (0.1%) |  |
| Education level |  |  |  |  |  |
| Primary or less | 157 (11.7%) | 35 (29.9%) | 101 (44.7%) | 21 (2.1%) | <0.001 |
| Secondary | 1034 (77.1%) | 54 (46.2%) | 79 (35.0%) | 901 (90.3%) |  |
| Tertiary | 139 (10.4%) | 28 (23.9%) | 35 (15.5%) | 76 (7.6%) |  |
| Key population identity characteristics |  |  |  |  |  |
| None reported | 247 (18.4%) | 98 (83.8%) | 149 (65.9%) | 0 (0.0%) | 0.004<sup>2</sup> |
| Any reported identity characteristic | 1083 (80.8%) | 19 (16.2%) | 66 (29.2%) | 998 (100.0%) |  |
| Ever injected drugs |  |  |  |  |  |
| No – never injected | 328 (24.5%) | 107 (91.5%) | 196 (86.7%) | 25 (2.5%) | <0.001 |
| Yes – injected drugs | 999 (74.5%) | 10 (8.5%) | 16 (7.1%) | 973 (97.5%) |  |
| HIV status |  |  |  |  |  |
| Negative | 521 (38.9%) | 0 (0.0%) | 44 (19.5%) | 477 (47.8%) | - |
| Positive | 616 (45.9%) | 18 (15.4%) | 156 | 442 (44.3%) |  |
|  |  |  | (69.0%) |  |  |
| Unknown | 204 (15.2%) | 99 (84.6%) | 26 (11.5%) | 79 (7.9%) |  |
| Testing characteristics |  |  |  |  |  |
| Assistance provided during self-testing |  |  |  |  |  |
| Assisted | 1164 (86.8%) | 31 (26.5%) | 135 (59.7%) | 998 (100.0%) | <0.001<sup>2</sup> |
| Unassisted | 166 (12.4%) | 86 (73.5%) | 80 (35.4%) | 0 (0.0%) |  |
| Test type |  |  |  |  |  |
| Blood | 708 (52.8%) | 70 (59.8%) | 131 (58.0%) | 507 (50.8%) | 0.042 |
| Oral | 633 (47.2%) | 47 (40.2%) | 95 (42.0%) | 491 (49.2%) |  |
| Facility type |  |  |  |  |  |
| HIV ART clinic | 164 (12.2%) | 18 (15.4%) | 146 (64.6%) | 0 (0.0%) | - |
| Community-based organisation (CBO) | 1,098 (81.9%) | 20 (17.1%) | 80 (35.4%) | 998 (100.0%) |  |
| Outpatient department (OPD) | 79 (5.9%) | 79 (67.5%) | 0 (0.0%) | 0 (0.0%) |  |

\*Missingness: Age group n=11, Education level n=11, Ever injected drugs n=14, Key Population identity n=11, Assistance n=11 (all from Nigeria).
<sup>†</sup> Chi-square test, if empty conditions not met for test (<5 in any cell). Where empty conditions are driven solely because of the singularity of the South African country context, comparisons are made between Cameroon and Nigeria only and specified in subsequent footnotes.
<sup>1</sup>Category ≥55 collapsed into ≥45 to fulfil requirements for chi-square test
<sup>2</sup>Key population identity chi square test between Cameroon and Nigeria only, as South African data had 0 participants with no reported identity characteristics.
<sup>3</sup>Assistance provided chi square test between Cameroon and Nigeria only, as South African data had 0 participants testing unassisted.

Participants were oldest in Cameroon, where just under three-quarters of people testing HCV positive were 55 years and above (71.8%, 84/117). In Nigeria, ages were more evenly spaced with three quarters of participants falling relatively equally between the ages of 25 and 54 (77.5%, 176/227). In South Africa, participants were much younger with over two-thirds of participants between the ages of 25 years and 34 years old (69.3%, 692/998). There were also differences by gender; in Cameroon, the gender distribution was relatively balanced with slightly more females testing positive (53.8%, 63/117), compared to two thirds of female participants in Nigeria (61.5%, 139/226) and almost all male participants in South Africa (92.3%, 921/998). Key population identity characteristics also varied by country: In Cameroon, over two-thirds of participants expressed no key population identity characteristics (83.8%, 98/117) and most had never injected drugs (91.5%, 107/117). Key population identity while not substantial in Nigeria, was greater than Cameroon, with just under a third of participants reporting a key population identity characteristic (29.2%, 66/226) although most also never having injected drugs (86.7%, 196/226). In South Africa, reported drug use (and consequently key population identity) was near universal (97.5%, 973/998). Finally, in Cameroon, participants were unlikely to be living with HIV, most participants had an unknown HIV status, with the minority reported as living with HIV because they had accessed testing through ART services (15.4%, 18/117). Nigeria, however, had the greatest burden of HIV, where more than two-thirds of participants with positive HCV were also living with HIV (69.0%, 156/226), followed by South Africa, where almost half of participants were HCV/HIV co-infected (44.3%, 442/998.)

### Successful HCV cascade progression

Cameroon had the greatest proportion of treatment initiation amongst those with PCR-confirmed viraemia (98.6%, 71/72) followed by Nigeria (90.8%, 168/185). In South Africa, only a small proportion of participants with detectable viraemia initiated treatment (4.3%, 37/854) (**Figure 1**).

**Figure 1.**
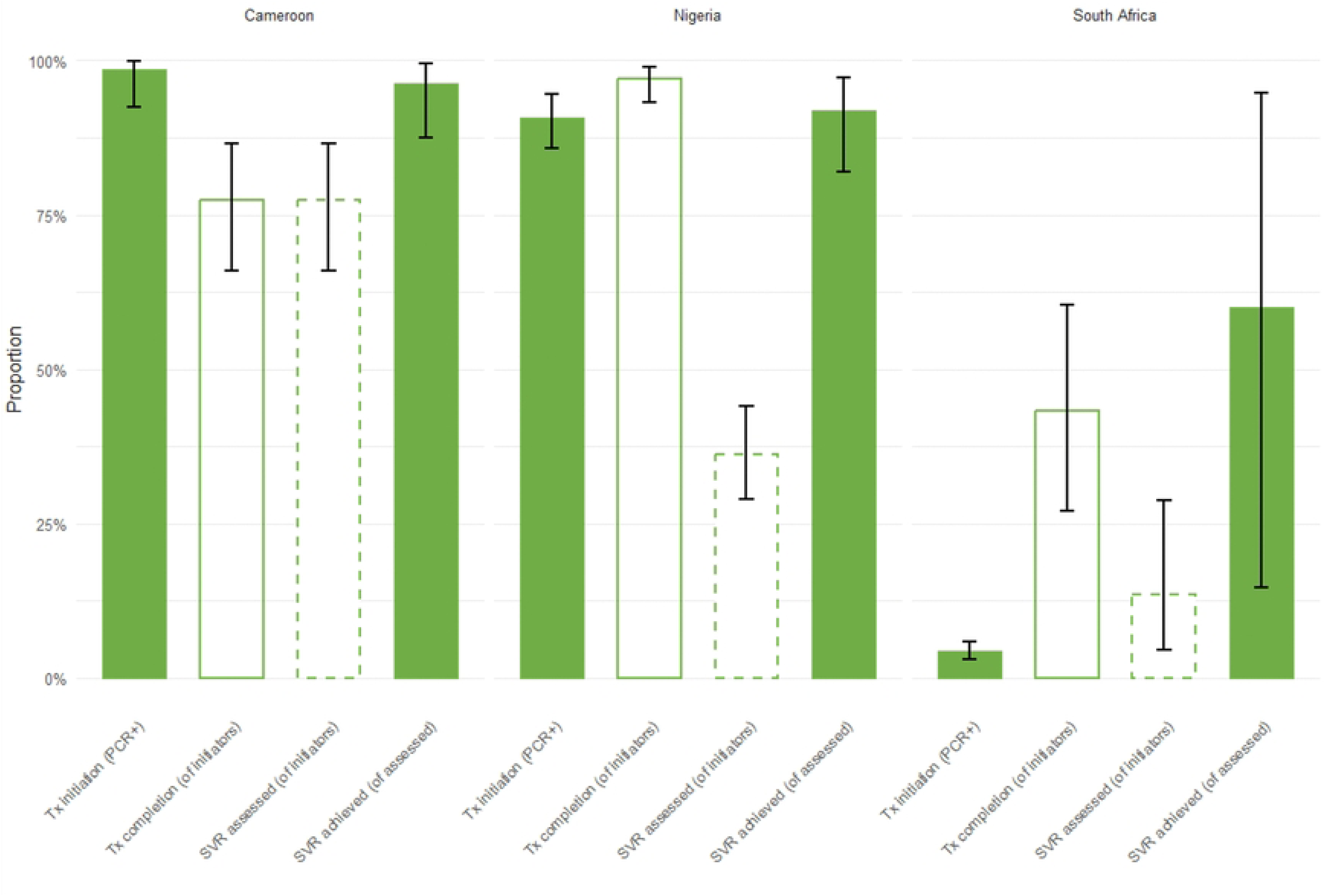
Proportions and binomial exact confidence intervals of completed cascade steps by country, N=1341 (Cameroon, n=117, Nigeria n=226, South Africa n=998); solid green bars are WHO defined cascade progression outcomes, i.e., treatment initiation and SVR. Dashed green is WHO defined additional indicators [30].

SVR among participants with a documented treatment outcome by the end of the reporting period was near universal in Cameroon; SVR among those with a recorded SVR test was near universal; almost all participants tested achieved SVR (96.4%, 53/55), and every participant that completed treatment tested for SVR. Of the 71 who initiated treatment in Cameroon, just over three-quarters completed treatment (77.5%, 55/71) with all these participants undergoing SVR testing. In Nigeria, cure among those tested was also high (91.8%, 56/62). However, SVR testing coverage was low with just over a third undergoing SVR testing (37.4%, 61/163). Conversely however, in Nigeria, treatment completion was near universal once therapy was initiated (97.0%, 163/168). In South Africa, SVR among those tested was lower; just under two-thirds of recorded SVR outcomes resulted in cure (60.6%, 3/5). Treatment completion was also limited at less than half (43.2%, 16/37) and less than a third of participants who completed treatment undergoing SVR testing (31.2%, 5/15).

### Cumulative incidence of disengagement

Cameroon, Nigeria and South Africa showed substantially different cumulative incidence of disengagement (**Table 3**).

**Table 3.** Model-based cumulative incidence of disengagement by country, at key timepoints (4, 12, 24 weeks and end of observation period)^1,2^.

| Country | 4 weeks (95% CrI) | 12 weeks (95% CrI) | 24 weeks (95% CrI) | End of observation period (95% CrI) |
| --- | --- | --- | --- | --- |
| *Cameroon* | 18.8% (15.6% to 23.4%) | 33.9% (28.3% to 41.1%) | 45.2% (37.8% to 53.6%) | 52.4% (44.4% to 61.2%) |
| *Nigeria* | 7.3% (5.3% to 9.9%) | 14.2% (10.6% to 18.4%) | 20.6% (15.9% to 26.0%) | 24.4% (19.1% to 30.3%) |
| *South Africa* | 74.6% (73.1% to 76.5%) | 77.7% (76.0% to 79.6%) | 77.9% (76.2% to 79.8%) | 77.9% (76.2% to 79.8%) |

<sup>1</sup>Estimates were obtained from posterior predictive cumulative incidence functions derived from cause-specific Weibull competing-risks models, evaluated at 4, 12, and 24 weeks (28, 84, and 168 days) and at the country-specific end of observation. End-of-observation estimates reflect differing follow-up durations: 272 days in Cameroon, 239 days in Nigeria, and 329 days in South Africa.
<sup>2</sup>The pooled Cameroon–Nigeria model excluded 11 participants due to missing time-to-event information (analytic N=329); no exclusions occurred in the South African model (N=998).

In Cameroon, the cumulative incidence of disengagement was just over half by the end of the observation period (52.4%; 95% CrI 44.4% to 61.2%), gradually increasing over time having reached 18.8% (95% CrI 15.6% to 23.4%) by the first month (4-weeks). Nigeria had the lowest cumulative incidence of disengagement over the follow-up period (24.4%; 95% CrI 19.1% to 30.3%) with low disengagement by the first month post-HCV self-testing (7.3%; 95% CrI 5.3% to 9.9%).

Disengagement was the highest in South Africa, over three-quarters cumulative incidence of disengagement over follow-up (77.9%; 95% CrI 76.2% to 79.8%) almost all of which had occurred by the first month (74.6%; 95% CrI 73.1% to 76.5%). Cumulative incidence plots reinforced these country-level differences in disengagement. In South Africa, almost all disengagement occurred almost immediately following HCV self-testing, reflecting minimal progression to treatment. In Cameroon and Nigeria, disengagement increased gradually over time, flattening earlier in Nigeria (**Fig 2**).

**Figure 2.**
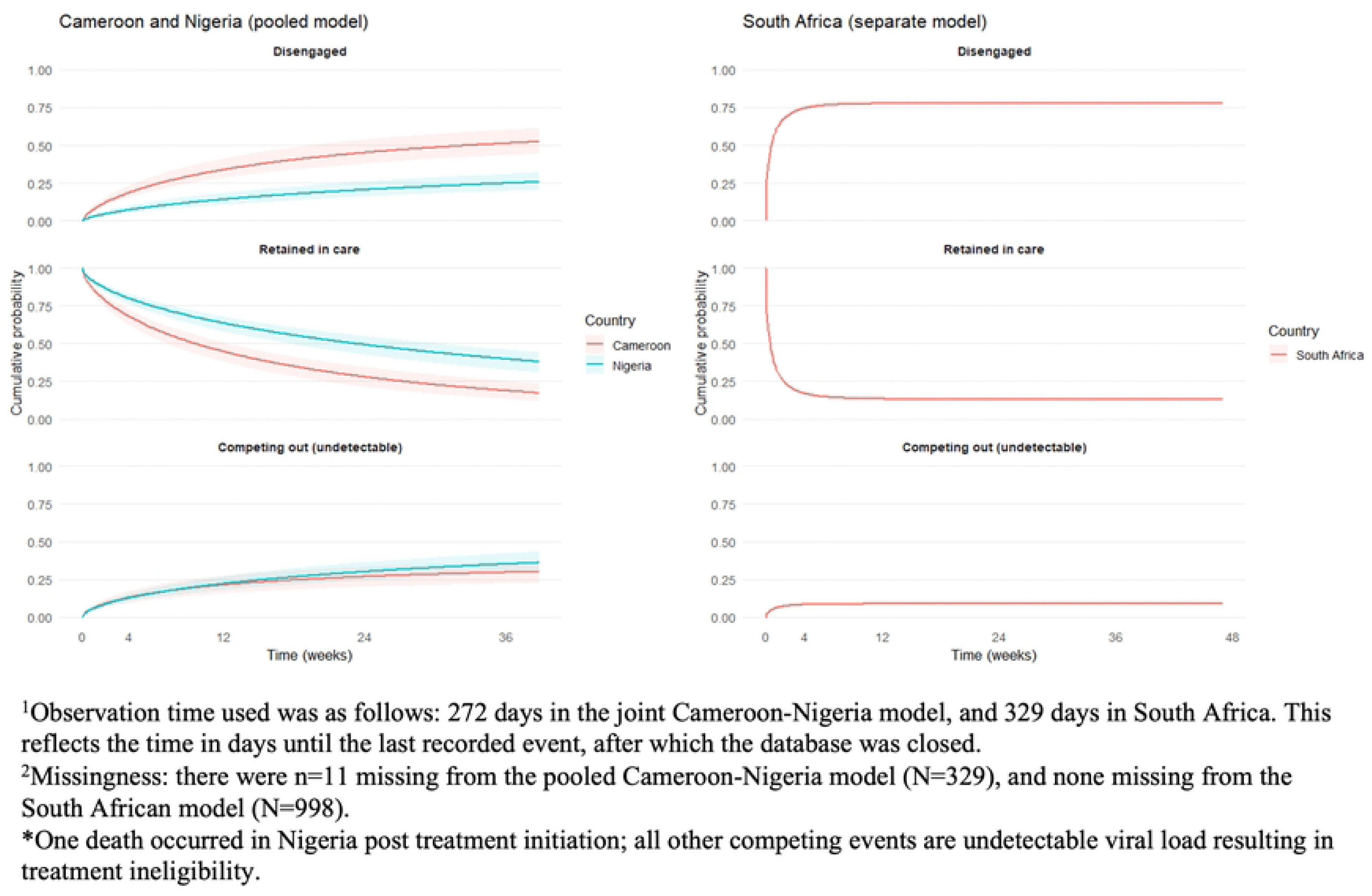
Model-based cumulative incidence of disengagement by country over the observation period, accounting for competing risks^1,2^.

However, after adjusting for site-level clustering in the Cameroon-Nigeria model, country-level fixed-effects were attenuated (HR: 1.00 95% CrI 0.68 to 1.49). All other fixed-effect covariate hazard ratios in the Cameroon-Nigerian data also showed minimal association with disengagement. Site-level heterogeneity (expressed as the site-level standard deviation of the random intercept on the log-time scale) in the Cameroon-Nigerian data dominated the model at 9.70 (95% CrI 6.17 to 15.47). The dominant source of systematic variation in disengagement appeared to arise from differences between sites rather than individual demographic or behavioural characteristics (**Table 4**).

**Table 4:** Adjusted hazard ratios with site-level random-effect estimates^1^.

| Covariate | Pooled Cameroon-Nigeria model (N=329*) | South African model (N=998) |
| --- | --- | --- |
|  | Adjusted HR (95% CrI) | Adjusted HR (95% CrI) |
| **Country** |  |  |
| Cameroon | - | - |
| Nigeria | 1.00 (0.68 to 1.49) | - |
| **Gender** |  |  |
| Female | - | - |
| Male | 0.90 (0.64 to 1.25) | 1.16 (0.92 to 1.46) |
| **Age** |  |  |
| 18-24 | - | - |
| 25-34 | 0.93 (0.66 to 1.33) | 1.01 (0.81 to 1.27) |
| 35-44 | 1.05 (0.73 to 1.49) | 1.03 (0.81 to 1.32) |
| 45-54 | 1.09 (0.76 to 1.55) | 1.24 (0.79 to 1.91) |
| 55+ | 0.91 (0.63 to 1.32) | 1.02 (0.58 to 1.79) |
| **Education** |  |  |
| Primary or less | - | - |
| Secondary | 0.83 (0.60 to 1.14) | 1.04 (0.76 to 1.43) |
| Tertiary | 0.98 (0.70 to 1.37) | 1.18 (0.83 to 1.66) |
| **Test Type** |  |  |
| Blood-based | - | - |
| Oral-test | 1.11 (0.80 to 1.52) | 0.94 (0.82 to 1.07) |
| **Mode of assistance** |  |  |
| Assisted | - | - |
| Unassisted | 1.00 (0.72 to 1.41) | - |
| **Key population Identity** |  |  |
| None reported | - | - |
| Yes | 0.99 (0.68 to 1.44) | - |
| **HIV status** |  |  |
| HIV negative | - | - |
| HIV positive | - | 1.96 (1.71 to 2.23) |
| Unknown HIV status | - | 1.29 (1.01 to 1.63) |
| **Site-level Standard Deviation** | **9.70 (6.17 to 15.47)** | **0.65 (0.25 to 1.47)** |

\*Missingness: there were n=11 missing from the pooled Cameroon-Nigeria model (N=329), and none missing from the South African model (N=998).

In South Africa, however, given the overall high rate of disengagement, sites were far more homogenous with a much smaller site-level standard deviation of the random intercept of 0.65 (95% CrI 0.25 to 1.47). Covariate-adjusted hazard ratios, unlike the Cameroon-Nigeria model, also showed tendencies toward differences in disengagement by demographic and clinical characteristics. Male sex (HR: 1.16 95% CrI 0.92 to 1.46), being aged between 45 and 54 years old (HR: 1.24, 95% CrI 0.79 to 1.91), tertiary education (HR: 1.18, 95% CrI 0.83 to 1.66) were all suggestive of faster disengagement. The most notable effect size in South Africa was HIV positive status, associated with an increase in the hazard of disengagement of almost two-fold compared to HIV-negative participants (HR 1.96; 95% CrI 1.71 to 2.23). An unknown HIV status was also associated with an increased hazard of disengagement of almost a third (HR: 1.29, 95% CrI 1.01 to 1.63).

Inspection of the site-specific posterior predictive probability of disengagement at 24-weeks derived from the Cameroon-Nigeria model confirmed substantial variation across sites. For a typical participant (Female, 55 years and above, secondary education, assisted testing, blood-based HCV self-test, and no reported key population identity), the posterior predictive probability of disengagement ranged by site from 1.7% (95% CrI 0.1 to 5.3%; OLA, Nigeria) to 75.3% (95% CrI 50.5 to 89.8%; YDF, Cameroon) (**Fig 3**).

**Figure 3.**
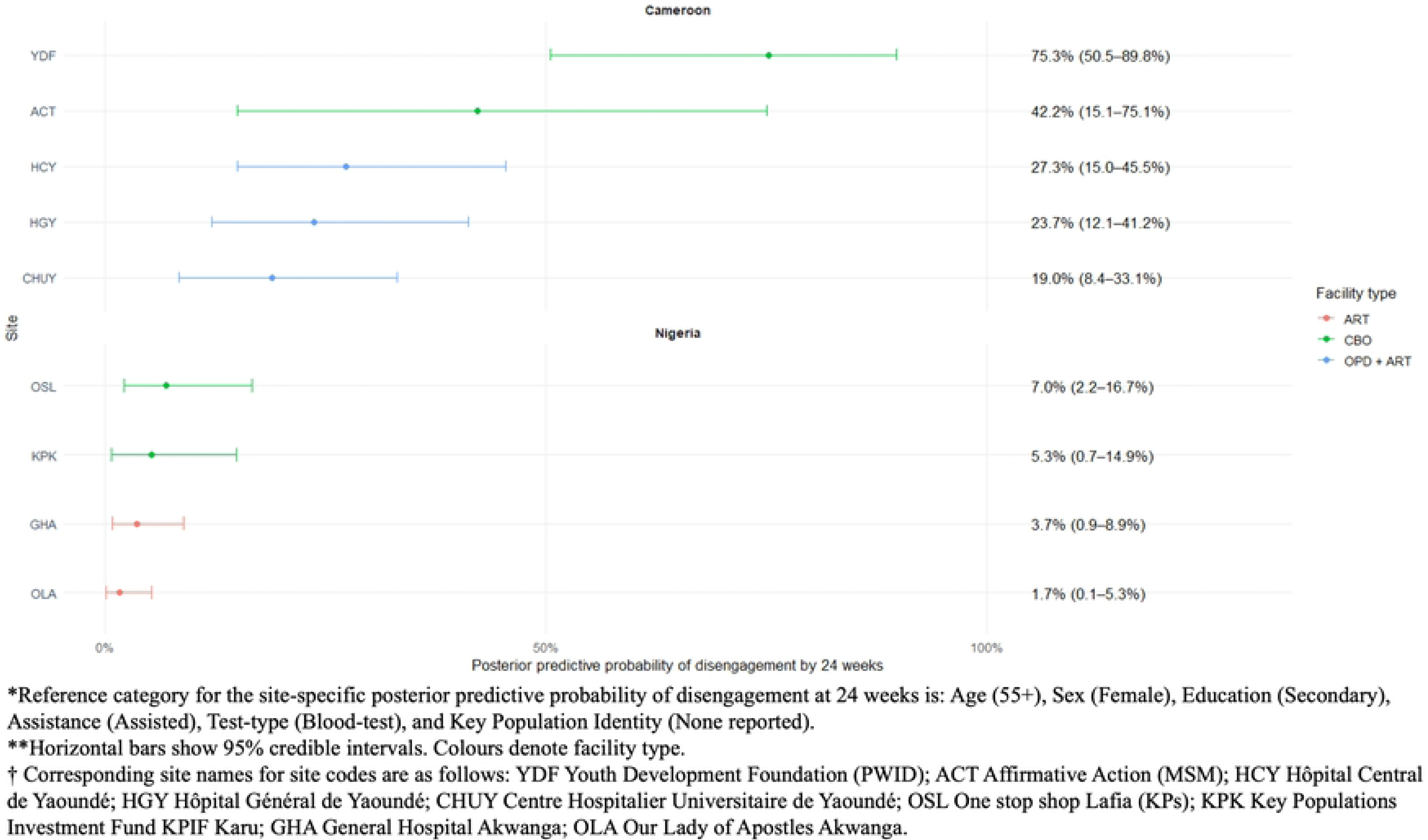
Site-specific posterior predictive probability of disengagement at 24-weeks (Cameroon and Nigeria dataset, N=329).

Visual stratification by country and facility type highlighted consistent differences in the predicted probability of disengagement by 24-weeks. both Cameroon and Nigeria, sites operating within hospital specialist services (ART clinics or combined OPD + ART services) showed lower posterior predictive probabilities of disengagement for the reference participant profile (Cameroon range 19.0% to 27.3%; Nigeria range 1.7% to 3.7%). In contrast, CBO sites demonstrated substantially higher predicted disengagement (Cameroon 42.2% to 75.3%; Nigeria 5.3% to 7.0%). These patterns suggest a facility-level gradient in retention, with CBOs showing markedly greater early disengagement than facility-based ART or mixed OPD/ART services.

### Disease severity outcomes

The greatest proportion of participants with viraemia was in South Africa (85.6%, 854/998), followed by three quarters in Nigeria (77.9%, 176/226) and just under two-thirds viraemic in Cameroon (61.5%, 72/117). Of those with detectable viral load, median viral loads were higher in Cameroon (1.15 × 10⁶ IU/mL, IQR 396 066–3.06 × 10⁶) than in Nigeria (2.24 × 10⁵ IU/mL, IQR 19 064–1.09 × 10⁶) (**Table 5**).

**Table 5.**
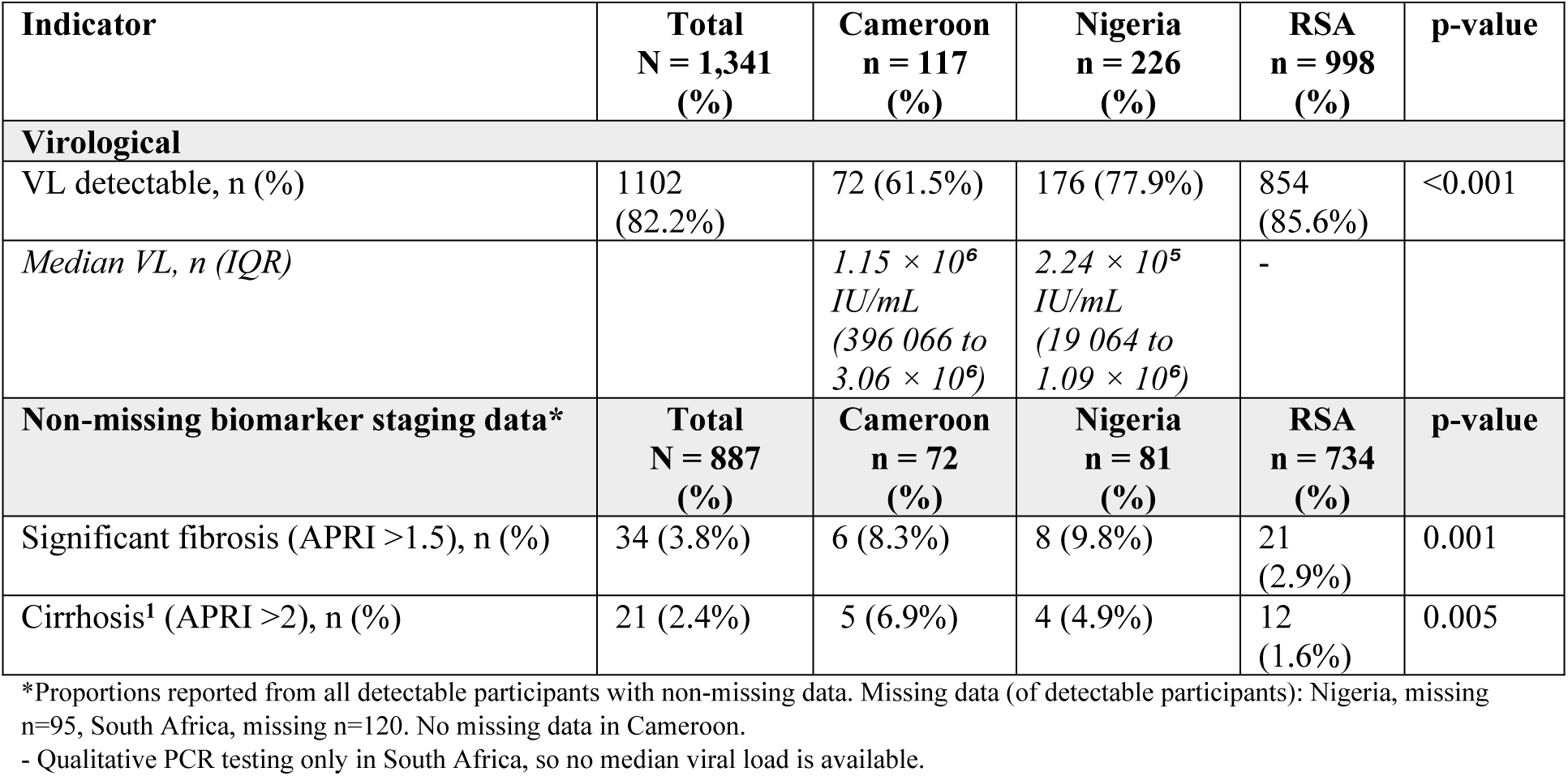
Disease severity profiles by country.

However, of those with chronic HCV infection in Cameroon and Nigeria, disease severity was greater. Although absolute participant numbers were small, more Cameroonian participants had significant fibrosis (8.3%, 6/72) of which a greater proportion was likely cirrhosis under WHO staging (6.9%, 5/72), compared to South Africa (fibrosis: 2.9%, 21/734, and cirrhosis 1.6%, 12/734). In Nigeria, of those with detectable disease and non-missing data, disease severity was also greater than South Africa (fibrosis: 9.8%, 8/81, and cirrhosis 4.9%, 4/81), although with extensive missing data (missing 54.0%, 95/176).

## Discussion

Despite comparable self-testing products and harmonised cascade definitions, there were substantial differences in disengagement from care across three STAR HCV-self-testing implementation studies (Cameroon, Nigeria and South Africa). Participants in Cameroon had the greatest treatment initiation and SVR, followed closely by Nigeria. In South Africa, treatment initiation was very low – with subsequent drop offs in all cascade outcomes. Crude country analyses demonstrated that disengagement from care was the lowest in Nigeria, reflecting the contextual differences in treatment initiation, and retention through to treatment completion and cure. In South Africa, a broadly homogenous implementation community-based delivery context, where disengagement was incredibly high, individual covariates suggestive of even greater disengagement were male gender, and HIV positive or unknown HIV status. HCV self-testing was able to be integrated into existing decentralized implementation efforts; however, the implementation context was key for disengagement outcomes; broadly Nigeria and Cameroon had lower disengagement than South Africa, and facility-based sites (specialist hospital services) had lower disengagement than community-based provision.

These differences in disengagement across contexts reflect key implementation differences by country. In the South African context, the delay in the country in the provision of DAAs (December 2023), and the initiation of a treatment protocol that prioritised DAAs for non-complex patients is likely partly responsible for the very low treatment initiation numbers seen in this context. In addition, South Africa had a young male population whose primary risk factor was injecting drug use. There is extensive literature on the challenges experienced by PWID in accessing centralised health services [13, 37, 38]. That HIV status was associated with greater disengagement in this context is plausible; in an already highly vulnerable population, HIV/HCV coinfection may be a marker of additional compounded vulnerability [39] and consequent structural barriers to care. For those who were not aware of their status, given that this service was integrated into an existing harm reduction service with high rates of HIV testing, this may be a sub-group generally less likely to engage in health services. Generally, in South Africa, despite a flexible and integrated HCV testing strategy within mobile van outreach and existing harm reduction services, aligning with WHO best practice [12], the delay in DAA provision, centralised treatment initiations and requirement for Directly Observed Therapy for treatment may all have presented structural barriers to care.

In Cameroon and Nigeria, implementation context was the primary indicator of disengagement; at the country-level, a key implementation difference between the Cameroon and Nigeria was the use of intensive follow-up and supported treatment referral in Nigeria compared to a less active process in Cameroon. Evidence from the HIV self-testing literature on the barriers to care engagement identified active follow-up as potentially supporting engagement in care [13]. WHO guidelines also endorse peer or lay support to accompany testing with navigation and follow-up [12]. However, it is important not to over-attribute this to country-level programming - Nasarawa is one of the only states in Nigeria benefitting from decentralised HCV RNA confirmation in each district, with one of the most mature viral hepatitis programmes in Nigeria [28]. Perhaps more importantly across both Cameroon and Nigeria were the descriptive difference in disengagement in ART and OPD services than community-based services. It may be that facility-models have clearer care pathways, especially for more treatment experienced participants. Integrating HCV self-testing in ART and OPD services is “low hanging fruit [28]” for HCV self-testing integration. However, HCV self-testing is being promoted as a complement to facility-based testing in decentralised settings such as community-based harm reduction services [30]; in these contexts, although decentralised models of HIV treatment are established [40], they may not yet be optimised for HCV treatment which continues to rely on referral to existing statutory or specialist services for care [41–43]. Further optimising HCV community care pathways is likely fundamental to address this gap.

Finally, the differences in populations testing positive also highlights a key study finding about each county programme’s success in reaching those most in need of treatment. In each country programme, HCV self-testing reached people at risk of HCV who were reflective of the background epidemic, and who had levels of viraemia requiring treatment that surpass existing global and regional estimates [3, 5, 44]. A clear minority were also experiencing severe disease, putting them at risk of liver failure or progression to hepatocellular carcinoma. The most severe disease was found in Cameroon, where participants were older. Although participants had less detectable viral loads (61.5%), suggestive of more cases of resolved past infection, those with detectable HCV had as higher proportion of liver scarring, inferring these participants had likely lived with the disease for many years especially when compared to the very young South African participant cohort with high levels of viraemic HCV but far less markers of advanced disease [5]. In Nigeria, higher proportions of active viraemia and scarring may reflect the greater proportion of Nigerian participants living with HIV, for whom clearance is a challenge, as HIV co-infection is associated with reduced spontaneous clearance of HCV and accelerated liver fibrosis progression [45–47]. However, large amounts of missing data in the Nigerian programme limit any concrete conclusions.

This study was conducted in three very different epidemic contexts in Africa, which aligned with WHO guidelines on optimised HCV delivery models to introduce integrated HCV ST into existing services [12]. However, one of the key study limitations is that the level of difference between contexts was not sufficiently captured through quantitative analyses. Survival analyses identified high site-level heterogeneity in pooled analyses. Because sites were fully nested within only two countries; the level of heterogeneity identified reflects both facility- and country-level effects. Any further conclusions as to the specifics of the implementation differences discussed above remain speculative without the benefit of provider and participant interview. Although implementers are authors on this study, capturing implementer and participant perspectives as part of embedded research design going forward would strengthen future conclusions [48], especially on participants reasons for disengagement.

We also abstracted from clinical data for treatment initiation and outcomes, with which there are known challenges with missing data. To mitigate this, we took a punitive approach to missing data in our analyses, where we assumed those who continued their engagement were more likely to have their data recorded programmatically, therefore missing data was coded as the outcome not happening. As treatment outcome data was abstracted within the set study period, individuals who were adherent but due to complete treatment after study end, would be falsely classed as disengaged. In turn, if people did not initiate treatment within the study period, they would be classed as disengaged, even if they later engage for care. The HIV self-test literature has shown that although most people engage for care within a week or month post-test, some may take more time to engage [49–51]. Conversely, participant recruitment was pragmatic making self-selection within service-attending populations likely. Individuals who accept HCVST may be more willing to engage in testing and study procedures and may be more motivated or better positioned to navigate linkage pathways than other service users. Observed cascade progression may overestimate progression and retention. These are limitations that we clearly acknowledge.

The interpretation of elevated APRI scores as proxy indicators of fibrosis and cirrhosis attributable to HCV infection is another limitation. Alternative causes or contributors of APRI elevation such as alcohol use, metabolic liver disease, viral co-infections, and non-hepatic thrombocytopenia were not excluded and may reflect underlying comorbidities particularly among older participants and especially among those attending care for chronic diseases in Cameroon.

Finally, by the end of the implementation period across all three countries, HCVST had been incorporated into national hepatitis policy, with OraSure receiving national regulatory authority market authorisation. However, at the time of writing, continued implementation has been constrained by funding limitations, including gaps in PEPFAR and Global Fund support. As a result, HCVST is not currently being procured or routinely offered at the participating study sites. These findings reflect programme performance under supported implementation conditions rather than sustained national scale-up.

## Conclusion

HCV self-testing can be used to reach and diagnose diverse participants, be they older people in Cameroon, women living with HIV in Nigeria, or PWID in South Africa, as reflective of these highly different epidemic contexts. From a programmatic perspective, the integration of HCV self-testing models into existing services, whether they are chronic disease clinics, harm reduction services, or key population community-based initiatives represent a key study finding for other countries; HCV self-testing can be integrated into existing decentralized implementation efforts for a range of other health services to take advantage of these “low hanging fruit [28]”. However, for this integration to be successful, HCV treatment pathways also need to be optimised to enable ongoing engagement along the HCV care cascade and broader epidemic impact; low-threshold testing provision needs to be matched by low-threshold pathways of care (e.g., onsite confirmatory testing and same-day treatment initiation). Centralised DAA provision may result in situations where people diagnosed through HCVST cannot access timely treatment. This is especially pertinent in contexts of overburdened health systems and nascent HCV control programmes, and outside of study contexts where access to generic DAA formulations remain underfunded by global donors and national governments [37].

## Data Availability

In supplementary material

## Supporting documents

**S1. List of ethics approvals obtained for individual studies and cross-country analyses**

**S2. Supporting data**

